# Machine Learning - Based Bleeding Risk Predictions in Atrial Fibrillation Patients on Direct Oral Anticoagulants

**DOI:** 10.1101/2024.05.27.24307985

**Authors:** Rahul Chaudhary, Mehdi Nourelahi, Floyd W. Thoma, Walid F. Gellad, Wei-Hsuan Lo-Ciganic, Kevin P. Bliden, Paul A. Gurbel, Matthew D. Neal, Sandeep K. Jain, Aditya Bhonsale, Suresh R. Mulukutla, Yanshan Wang, Matthew E. Harinstein, Samir Saba, Shyam Visweswaran

## Abstract

**Importance:** Accurately predicting major bleeding events in non-valvular atrial fibrillation (AF) patients on direct oral anticoagulants (DOACs) is crucial for personalized treatment and improving patient outcomes, especially with emerging alternatives like left atrial appendage closure devices. The left atrial appendage closure devices reduce stroke risk comparably but with significantly fewer non-procedural bleeding events.

**Objective:** To evaluate the performance of machine learning (ML) risk models in predicting clinically significant bleeding events requiring hospitalization and hemorrhagic stroke in non-valvular AF patients on DOACs compared to conventional bleeding risk scores (HAS-BLED, ORBIT, and ATRIA) at the index visit to a cardiologist for AF management.

**Design:** Prognostic modeling with retrospective cohort study design using electronic health record (EHR) data, with clinical follow-up at one-, two-, and five-years.

**Setting:** University of Pittsburgh Medical Center (UPMC) system.

**Participants:** 24,468 non-valvular AF patients aged ≥18 years treated with DOACs, excluding those with prior history of significant bleeding, other indications for DOACs, on warfarin or contraindicated to DOACs.

**Exposure(s):** DOAC therapy for non-valvular AF.

**Main Outcome(s) and Measure(s):** The primary endpoint was clinically significant bleeding requiring hospitalization within one year of index visit. The models incorporated demographic, clinical, and laboratory variables available in the EHR at the index visit.

**Results:** Among 24,468 patients, 553 (2.3%) had bleeding events within one year, 829 (3.5%) within two years, and 1,292 (5.8%) within five years of index visit. We evaluated multivariate logistic regression and ML models including random forest, classification trees, k-nearest neighbor, naive Bayes, and extreme gradient boosting (XGBoost) which modestly outperformed HAS-BLED, ATRIA, and ORBIT scores in predicting clinically significant bleeding at 1-year follow-up. The best performing model (random forest) showed area under the curve (AUC-ROC) 0.76 (0.70-0.81), G-Mean score of 0.67, net reclassification index 0.14 compared to 0.57 (0.50-0.63), G-Mean score of 0.57 for HASBLED score, p-value for difference <0.001. The ML models had improved performance compared to conventional risk across time-points of 2-year and 5-years and within the subgroup of hemorrhagic stroke. SHAP analysis identified novel risk factors including measures from body mass index, cholesterol profile, and insurance type beyond those used in conventional risk scores.

**Conclusions and Relevance:** Our findings demonstrate the superior performance of ML models compared to conventional bleeding risk scores and identify novel risk factors highlighting the potential for personalized bleeding risk assessment in AF patients on DOACs.

## Introduction

Non-valvular atrial fibrillation (AF) is the most prevalent cardiac arrhythmia, posing a significant public health challenge with an expected prevalence of 12.1 million in the United States by 2030 ^1,2^. Current guidelines recommend using the CHALDSL-VASc score to assess stroke risk and suggest direct oral anticoagulants (DOACs) for high-risk patients ^3^. Recent updates expand stroke prevention strategies to include patients with device-detected subclinical AF, thereby expanding DOAC indications ^4–6^. Despite advances, oral anticoagulation is associated with annual major bleeding rates of 2%-4%, with case fatality rates of 8%-15% ^3,7–10^.

Transcatheter left atrial appendage closure has emerged as a viable alternative for patients with non-valvular AF at high thromboembolic risk who are unsuitable for long-term oral anticoagulant use ^11^. These devices offer comparable efficacy in stroke risk reduction and have a marked decrease in non-procedural bleeding events (up to 46%) in carefully selected populations with high bleeding risk ^12^. With the advent of such alternatives to DOACs, identifying patients at high risk of bleeding on DOACs becomes crucial for early intervention, especially before experiencing a sentinel significant bleeding event.

This study focuses on AF patients managed by cardiologists, as they often handle more complex cases, comorbidities, and higher risk profiles, and play a key role in managing DOAC therapy. Conventional risk scores used for assessing bleeding risk on anticoagulation have limitations when applied to dynamic and heterogeneous real-world patient populations. Registry and clinical trial data often fail to capture the complexity of real-world scenarios, where clinical decisions must be made with incomplete information, such as missing data on the duration of AF before being seen by cardiologists for further management. Consequently, clinical guidelines have shifted away from relying solely on bleeding scores, highlighting the need for more comprehensive and accurate risk assessment tools beyond the HAS-BLED ^13^, ATRIA ^14^, and ORBIT ^15^ scores.

Previous studies have explored the use of machine learning (ML) models in predicting bleeding risk in AF patients, demonstrating improved performance over conventional scoring systems ^16,17^. However, these studies have limitations, such as relying on registry-based data that may not accurately represent the complexity and heterogeneity of real-world clinical practice, focusing on broader contexts like predicting bleeding risk from antithrombotic therapy in general patient populations, or concentrating only on AF subpopulations ^18,19^. Moreover, they lack direct comparison between ML models and multiple conventional risk scores, with most studies comparing ML models to only individual risk scores, such as HAS-BLED ^20^. Our study aims to address this knowledge gap by developing ML models based on real-world electronic health record (EHR) data available to cardiologists at the time of the index visit for AF management among patients on DOACs and evaluating their performance against the majority of conventional risk scores used in clinical practice, including HAS-BLED, ORBIT, and ATRIA scores.

We hypothesize that ML risk models, using EHR data, can improve bleeding risk prediction of a clinically significant major bleeding event necessitating hospitalization among AF patients treated with DOACs compared to conventional bleeding risk scores.

## Methods

### Study Cohort and Design

This prognostic modeling with retrospective cohort study design, approved by the University of Pittsburgh Institutional Review Board, identified patients aged ≥18 years with non-valvular AF treated with DOACs within the University of Pittsburgh Medical Center (UPMC) system between January 1, 2010, and November 30, 2022. By focusing on patients managed by cardiologists within a single healthcare system (UPMC), the study ensures more consistent and comprehensive data collection through the electronic health records (EHRs). This approach enhances the study’s relevance and applicability to clinical decision-making in cardiology practice, where cardiologists often make decisions about AF management and bleeding risk assessment based on the information available at the time of the patient’s visit. Patients with no follow-up data at 2-year or 5-year follow-up were excluded from analyses at these timepoints and censored after the index event. Patients were excluded if they had 1) another indication for DOAC (e.g., venous thromboembolism); 2) history of major bleeding event requiring hospitalization; 3) on warfarin; 4) received left atrial appendage closure device; and 5) contraindication to DOACs (e.g., mechanical heart valve) despite the presence of off-label use in clinical practice **(Figure 1)**. We estimated the clinically significant bleeding risk based on clinical data available at the index visit to a cardiologist for management of AF, with clinical follow-up at one, two, and five years. The study size of 24,468 patients was deemed sufficient to develop and validate the prediction models based on the expected event rate and the number of candidate predictors. The study adhered to the TRIPOD+AI guidelines ^21^.

**Figure 1.**
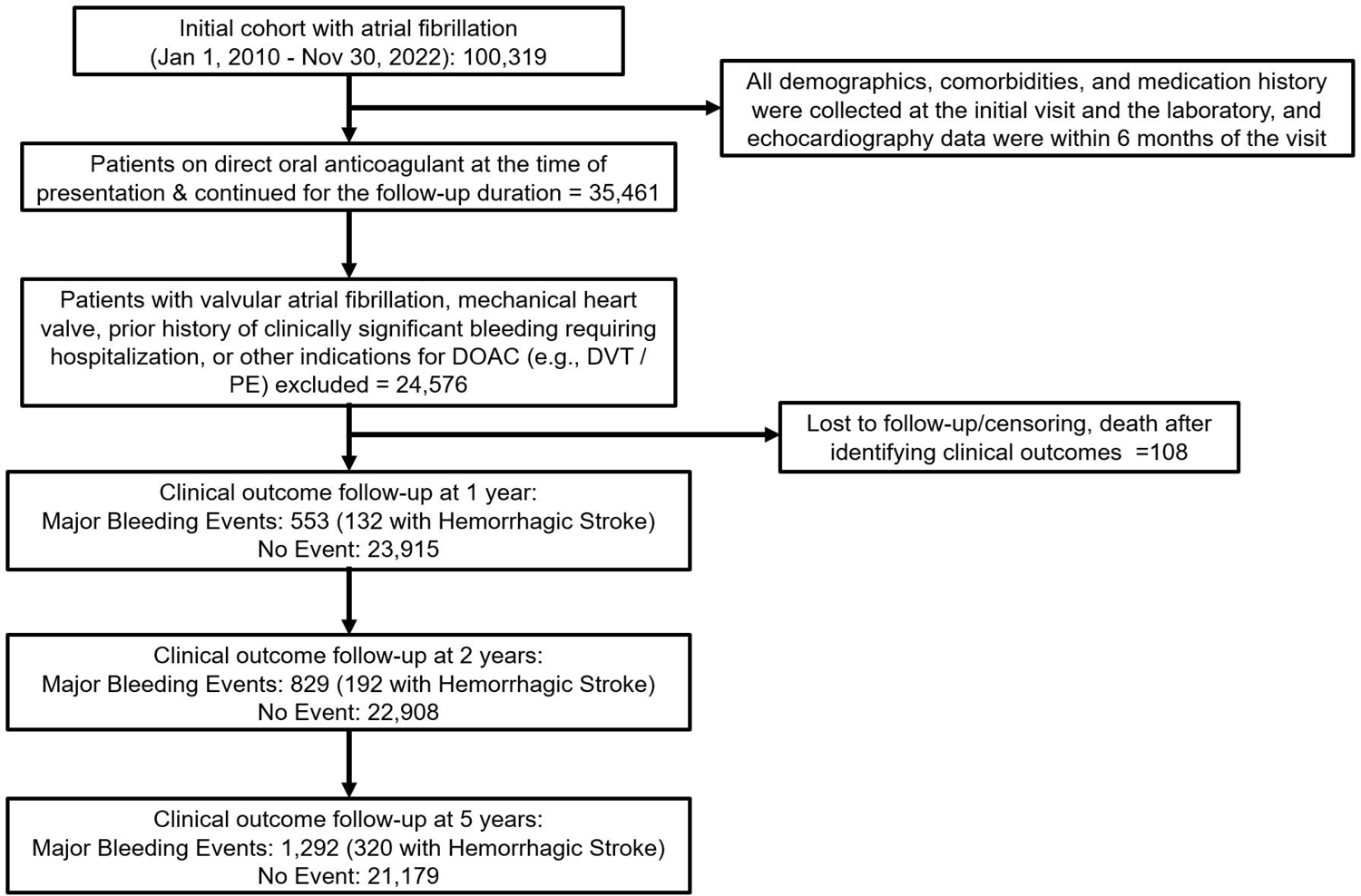
Study flow diagram.

### Data Collection and Outcomes

Clinical and demographic data were extracted from the EHR for patients with varying duration of AF, with laboratory and echocardiography parameters within 6 months from the index visit to the cardiologist. Follow-up lasted until November 30, 2022, or the first adverse event. Patients who died during the follow-up period without experiencing a bleeding event were censored at the time of death. The primary endpoint was an incident clinically significant bleeding event requiring a hospitalization (including gastrointestinal bleeding and hemorrhagic stroke) within one year since index visit, identified through validated administrative diagnosis codes (ICD-9 and ICD-10) from the EHR database. Patients were censored after the first bleeding event. The secondary endpoints included an incident bleeding event at two and five years of follow-up and incident hemorrhagic stroke at one, two and five years of follow-up **(Supplemental Table 1)** ^22,23^. These codes, listed as one of the top three diagnoses for an inpatient admission, captured significant bleeding events and hemorrhagic strokes, as detailed in **supplementary table 1**. The outcome assessment was not blinded, as the bleeding events were identified through validated administrative diagnosis codes from the EHR database.

### Candidate Predictors and Machine Learning

Data preparation involved discarding variables with over 60% missing values to avoid bias, and the remaining missing data were imputed with median values for continuous variables and mode values for categorical variables, after determining the data were missing completely at random (MCAR) using Little’s MCAR test ^24^ (**Supplementary Table 2)**. New clinically relevant variables were generated from data within 6 months of index visit through feature engineering, such as mean arterial pressure from systolic and diastolic blood pressure, prediabetes status from hemoglobin A1C and diabetes history, and poorly controlled hypertension from systolic blood pressure ^25^. Additionally, multicollinearity and variance inflation (VIF) were assessed ^26–28^. The final set of variables was selected based on recursive feature elimination ^29^, multicollinearity assessment, and domain expertise to ensure the inclusion of the most robust and informative features in model training. We applied commonly used ML algorithms, including multivariate logistic regression with L1 (Lasso) and L2 (Ridge) regularization, random forest, extreme gradient boosting (XGBoost), classification trees, k-nearest neighbor (KNN), and naïve Bayes ^26–28^. The selected ML models all had their strengths and limitations. While multivariate logistic regression with L1 and L2 regularization addressed potential overfitting and feature selection, random forest and XGBoost leveraged ensemble learning approaches ^30^. Classification trees delivered a transparent, rule-based classification method, and KNN and naïve Bayes were chosen for their proficiency in handling nonlinear data patterns ^31^. SHAP (SHapley Additive exPlanations) analysis was conducted for feature importance and model explainability ^32^. HAS-BLED, ORBIT, and ATRIA scores were computed for benchmarking.

### Training and Validation

In this study, we used a stratified splitting to partition the data into a training set (70%) and two test sets (15% dataset with low-comorbidities, and 15% random), ensuring that the proportion of female sex, and Black race were similar across the datasets to ensure generalizability of developed models. This technique is commonly employed with imbalanced datasets, where the minority class has considerably fewer instances than others to similar class distributions in the training and test sets ^33^. We employed two test sets: (1) a low comorbid population with a low Charlson comorbidity index (< 2) and no major bleeding history, and (2) a randomly selected test set mirroring the training data distribution. Different sampling techniques (under-sampling and over-sampling) and ratios (1:1, 1:5, 1:10, 1:20) were employed to address class imbalance and evaluate their impact on model performance. These techniques were chosen to ensure adequate representation of the minority class (adverse events) during model training and to assess the robustness of the ML models under different class distributions ^34^. A ten-fold stratified cross-validation approach was used to ensure robust validation of the model’s performance ^35^.

### Performance Measures and Statistical Analysis

We assessed model performance using the area under the receiver-operator characteristic curve (AUC-ROC) and area under the precision-recall curve (AUPRC). Additionally, we computed the net reclassification index (NRI) and integrated discrimination improvement (IDI) to evaluate the incremental value of the ML models compared to HASBLED score as baseline. We computed Brier score and log loss to quantify calibration ^36^. Calibration curves were also plotted to assess the agreement between predicted and observed probabilities. Risk stratification curves were plotted to visualize the distribution of predicted risk scores and their corresponding observed event rates. Youden’s index determined the optimal threshold for dichotomizing the model output and to calculate performance metrics, including sensitivity (recall), specificity, accuracy, precision (positive predictive value [PPV]), negative predictive value (NPV), F1 score (harmonic mean of the precision and recall scores) ^17^. Due to the anticipated significant class imbalance, we evaluated the overall performance of the algorithm using the G-Mean Score (geometric mean of sensitivity and specificity) ^37,38^. The G-mean score provides a balanced measure of performance, with higher values indicating better performance in correctly identifying both the majority and minority classes, making it particularly useful in imbalanced datasets.

We presented continuous variables as median and interquartile range, and categorical variables as frequencies and percentages. Mann-Whitney U test and Chi-square test were used for comparing continuous and categorical variables, respectively, between patients with and without an adverse event. The performance of HAS-BLED, ATRIA, and ORBIT risk scores was evaluated using identified thresholds for high bleeding risk (>2 for HAS-BLED, >3 for ORBIT, and >4 for ATRIA) ^39–41^. Additionally, low comorbidity and random test sets evaluated the model performance across diverse clinical scenarios, providing insights into model robustness. The SHAP analysis was performed for insights into the relative importance of features in the models’ decision-making processes ^42^. The magnitude and direction of the SHAP values indicate the average contribution of each feature to the model’s output, with positive and negative values signifying the impact on the prediction, thereby facilitating the interpretation of the models’ underlying logic. Statistical analyses were performed using the Python programming language version 3.12.2, with a two-sided p-value of <0.05 considered statistically significant and Bonferroni correction applied to mitigate the risk of Type I error across multiple comparisons ^43^.

## Results

### Baseline Characteristics

The study included 24,468 patients (median age 73.1 years, 45% women, 95.4% White) followed for up to five years since index visit. The incidence of bleeding events was 2.3%, 3.5%, and 5.8% at one, two, and five years, respectively. The incidence of hemorrhagic stroke was 0.5%, 0.8%, and 1.4% at the same time points. Patients who experienced a bleeding event at one year were older, higher proportion of Medicare or Medicaid insurance, higher comorbidity burden (including hypertension, prediabetes, coronary artery disease, heart failure, active cancer, anemia, peptic ulcer disease, and depression), aspirin use, and valvular diseases (moderate to severe mitral regurgitation, and severe aortic stenosis) at the time of index visit (p<0.01 individually for all) **(Table 1)**.

**Table 1.**
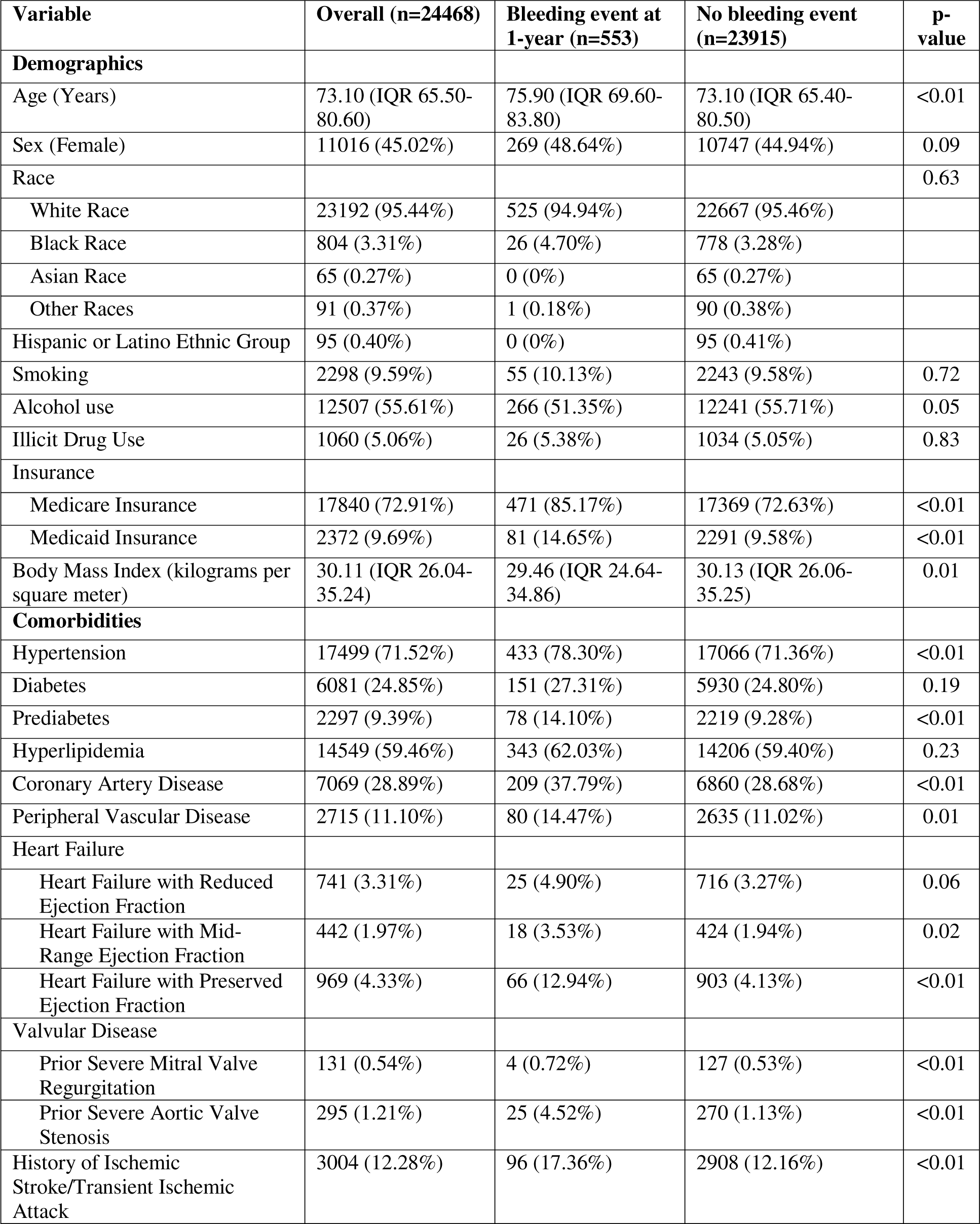
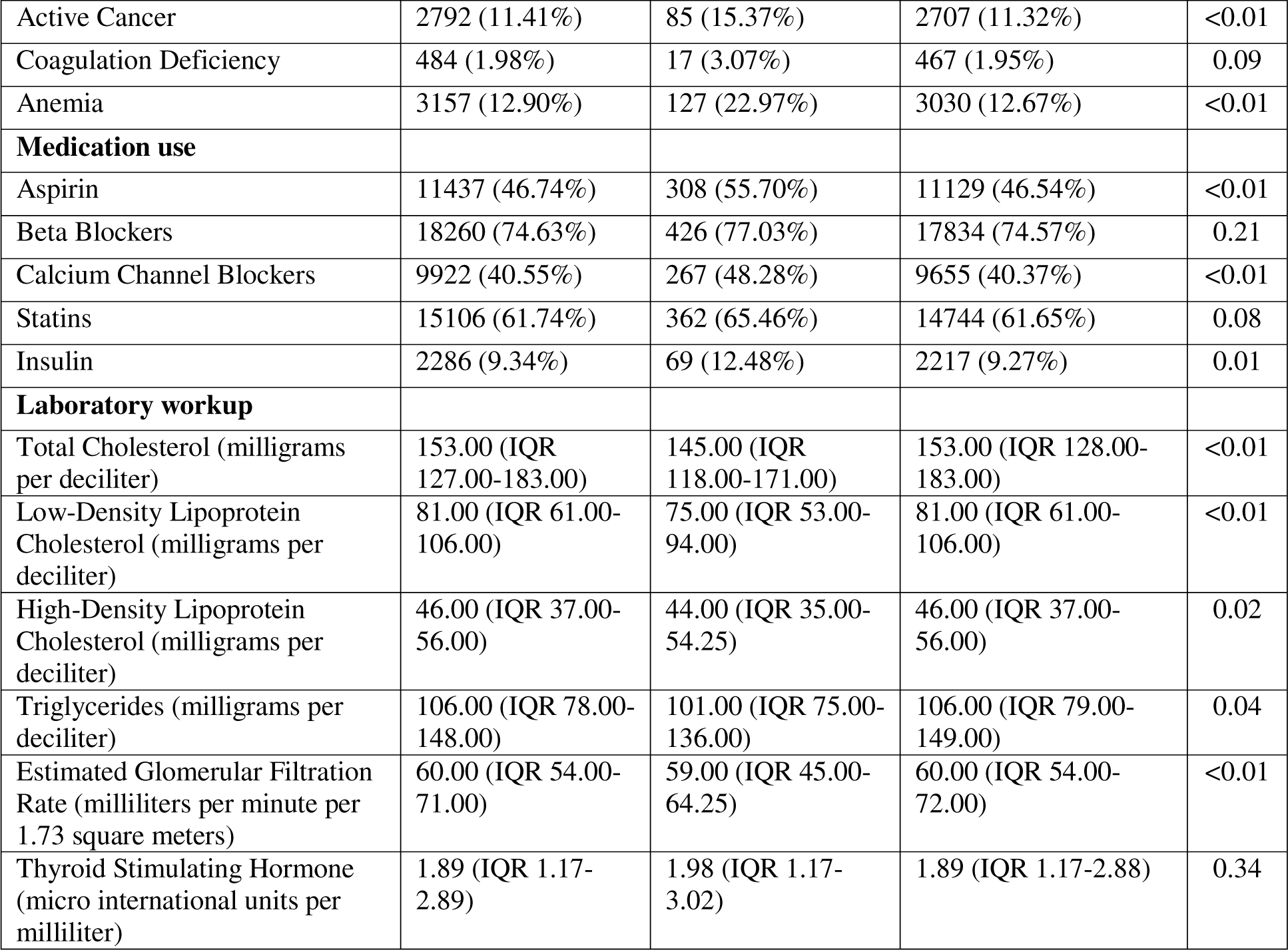
Characteristics of the patients at index date comparing patients who experienced major bleeding event at 1-year of follow-up versus without a major bleeding event at 1-year.

| Variable | Overall (n=24468) | Bleeding event at 1-year (n=553) | No bleeding event (n=23915) | p-value |
| --- | --- | --- | --- | --- |
| **Demographics** |  |  |  |  |
| Age (Years) | 73.10 (IQR 65.50-80.60) | 75.90 (IQR 69.60-83.80) | 73.10 (IQR 65.40-80.50) | <0.01 |
| Sex (Female) | 11016 (45.02%) | 269 (48.64%) | 10747 (44.94%) | 0.09 |
| Race |  |  |  | 0.63 |
| White Race | 23192 (95.44%) | 525 (94.94%) | 22667 (95.46%) |  |
| Black Race | 804 (3.31%) | 26 (4.70%) | 778 (3.28%) |  |
| Asian Race | 65 (0.27%) | 0 (0%) | 65 (0.27%) |  |
| Other Races | 91 (0.37%) | 1 (0.18%) | 90 (0.38%) |  |
| Hispanic or Latino Ethnic Group | 95 (0.40%) | 0 (0%) | 95 (0.41%) |  |
| Smoking | 2298 (9.59%) | 55 (10.13%) | 2243 (9.58%) | 0.72 |
| Alcohol use | 12507 (55.61%) | 266 (51.35%) | 12241 (55.71%) | 0.05 |
| Illicit Drug Use | 1060 (5.06%) | 26 (5.38%) | 1034 (5.05%) | 0.83 |
| Insurance |  |  |  |  |
| Medicare Insurance | 17840 (72.91%) | 471 (85.17%) | 17369 (72.63%) | <0.01 |
| Medicaid Insurance | 2372 (9.69%) | 81 (14.65%) | 2291 (9.58%) | <0.01 |
| Body Mass Index (kilograms per square meter) | 30.11 (IQR 26.04-35.24) | 29.46 (IQR 24.64-34.86) | 30.13 (IQR 26.06-35.25) | 0.01 |
| **Comorbidities** |  |  |  |  |
| Hypertension | 17499 (71.52%) | 433 (78.30%) | 17066 (71.36%) | <0.01 |
| Diabetes | 6081 (24.85%) | 151 (27.31%) | 5930 (24.80%) | 0.19 |
| Prediabetes | 2297 (9.39%) | 78 (14.10%) | 2219 (9.28%) | <0.01 |
| Hyperlipidemia | 14549 (59.46%) | 343 (62.03%) | 14206 (59.40%) | 0.23 |
| Coronary Artery Disease | 7069 (28.89%) | 209 (37.79%) | 6860 (28.68%) | <0.01 |
| Peripheral Vascular Disease | 2715 (11.10%) | 80 (14.47%) | 2635 (11.02%) | 0.01 |
| Heart Failure |  |  |  |  |
| Heart Failure with Reduced Ejection Fraction | 741 (3.31%) | 25 (4.90%) | 716 (3.27%) | 0.06 |
| Heart Failure with Mid-Range Ejection Fraction | 442 (1.97%) | 18 (3.53%) | 424 (1.94%) | 0.02 |
| Heart Failure with Preserved Ejection Fraction | 969 (4.33%) | 66 (12.94%) | 903 (4.13%) | <0.01 |
| Valvular Disease |  |  |  |  |
| Prior Severe Mitral Valve Regurgitation | 131 (0.54%) | 4 (0.72%) | 127 (0.53%) | <0.01 |
| Prior Severe Aortic Valve Stenosis | 295 (1.21%) | 25 (4.52%) | 270 (1.13%) | <0.01 |
| History of Ischemic Stroke/Transient Ischemic Attack | 3004 (12.28%) | 96 (17.36%) | 2908 (12.16%) | <0.01 |
| Active Cancer | 2792 (11.41%) | 85 (15.37%) | 2707 (11.32%) | <0.01 |
| Coagulation Deficiency | 484 (1.98%) | 17 (3.07%) | 467 (1.95%) | 0.09 |
| Anemia | 3157 (12.90%) | 127 (22.97%) | 3030 (12.67%) | <0.01 |
| **Medication use** |  |  |  |  |
| Aspirin | 11437 (46.74%) | 308 (55.70%) | 11129 (46.54%) | <0.01 |
| Beta Blockers | 18260 (74.63%) | 426 (77.03%) | 17834 (74.57%) | 0.21 |
| Calcium Channel Blockers | 9922 (40.55%) | 267 (48.28%) | 9655 (40.37%) | <0.01 |
| Statins | 15106 (61.74%) | 362 (65.46%) | 14744 (61.65%) | 0.08 |
| Insulin | 2286 (9.34%) | 69 (12.48%) | 2217 (9.27%) | 0.01 |
| **Laboratory workup** |  |  |  |  |
| Total Cholesterol (milligrams per deciliter) | 153.00 (IQR 127.00-183.00) | 145.00 (IQR 118.00-171.00) | 153.00 (IQR 128.00-183.00) | <0.01 |
| Low-Density Lipoprotein Cholesterol (milligrams per deciliter) | 81.00 (IQR 61.00-106.00) | 75.00 (IQR 53.00-94.00) | 81.00 (IQR 61.00-106.00) | <0.01 |
| High-Density Lipoprotein Cholesterol (milligrams per deciliter) | 46.00 (IQR 37.00-56.00) | 44.00 (IQR 35.00-54.25) | 46.00 (IQR 37.00-56.00) | 0.02 |
| Triglycerides (milligrams per deciliter) | 106.00 (IQR 78.00-148.00) | 101.00 (IQR 75.00-136.00) | 106.00 (IQR 79.00-149.00) | 0.04 |
| Estimated Glomerular Filtration Rate (milliliters per minute per 1.73 square meters) | 60.00 (IQR 54.00-71.00) | 59.00 (IQR 45.00-64.25) | 60.00 (IQR 54.00-72.00) | <0.01 |
| Thyroid Stimulating Hormone (micro international units per milliliter) | 1.89 (IQR 1.17-2.89) | 1.98 (IQR 1.17-3.02) | 1.89 (IQR 1.17-2.88) | 0.34 |

The baseline characteristics of the training and random test dataset were similar, with a median age of 73, 45% women, 3,4% Black race, and incident bleeding rate of 2.3% at 1-year of follow-up. The low-comorbidity test set had similar age, sex, and Black race but lower comorbidity burden and incident bleeding rate of 1.9% at 1-year of follow-up.

### Predictive Performance of Machine Learning Models

In both the low-comorbidity test and random test cohorts, ML risk models demonstrated superior performance in discriminative power, overall performance, risk stratification, and calibration compared to conventional risk scores (HAS-BLED, ORBIT, and ATRIA) in predicting bleeding events at the 1-year follow-up **(Table 2, Supplementary Table 3)**. In the low-comorbidity test cohort, the best performing ML model, XGBoost with AUC 0.69 (95% CI 0.63–0.74; G-Mean score 0.59; NRI 0.11; IDI 0.04, Brier score 0.04, log loss 0.19), outperformed the best performing conventional score HASBLED (AUC 0.54, 0.48-0.60; G-Mean score 0.53, Brier score 0.32, log loss 11.47), with p-values <0.001 for the individual differences **(Figure 2**, **Table 2, Supplementary Table 3)**.

**Figure 2.**
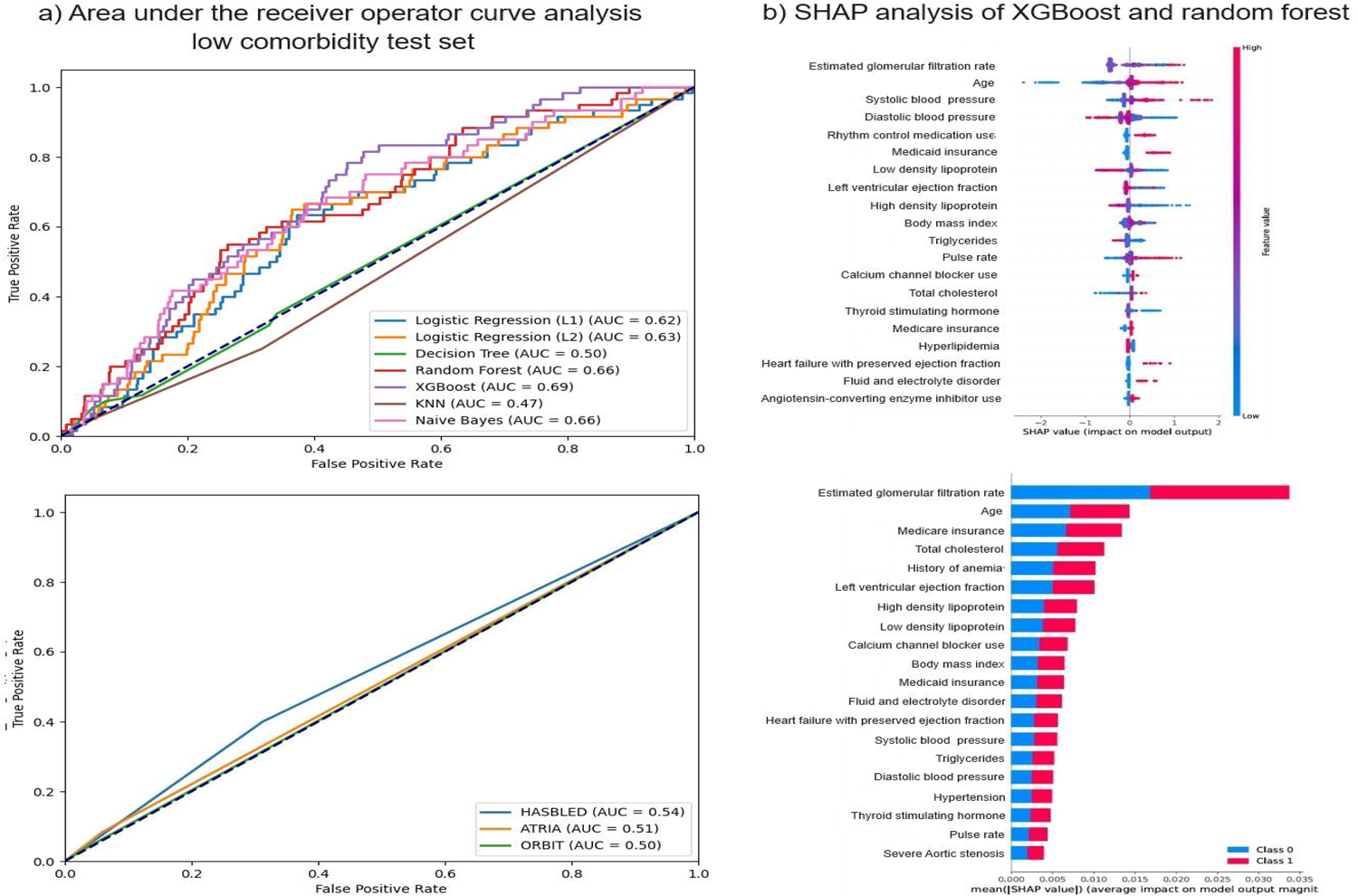
Comparative performance of machine learning models with HASBLED, ORBIT, and ATRIA scores in the low-comorbidity test set to predict a significant bleeding event at 1-year a) AUC-ROC analysis; and b) SHAP analysis for random forest model and extreme gradient boosting (XGBoost) model showing representative factors.

**Table 2.**
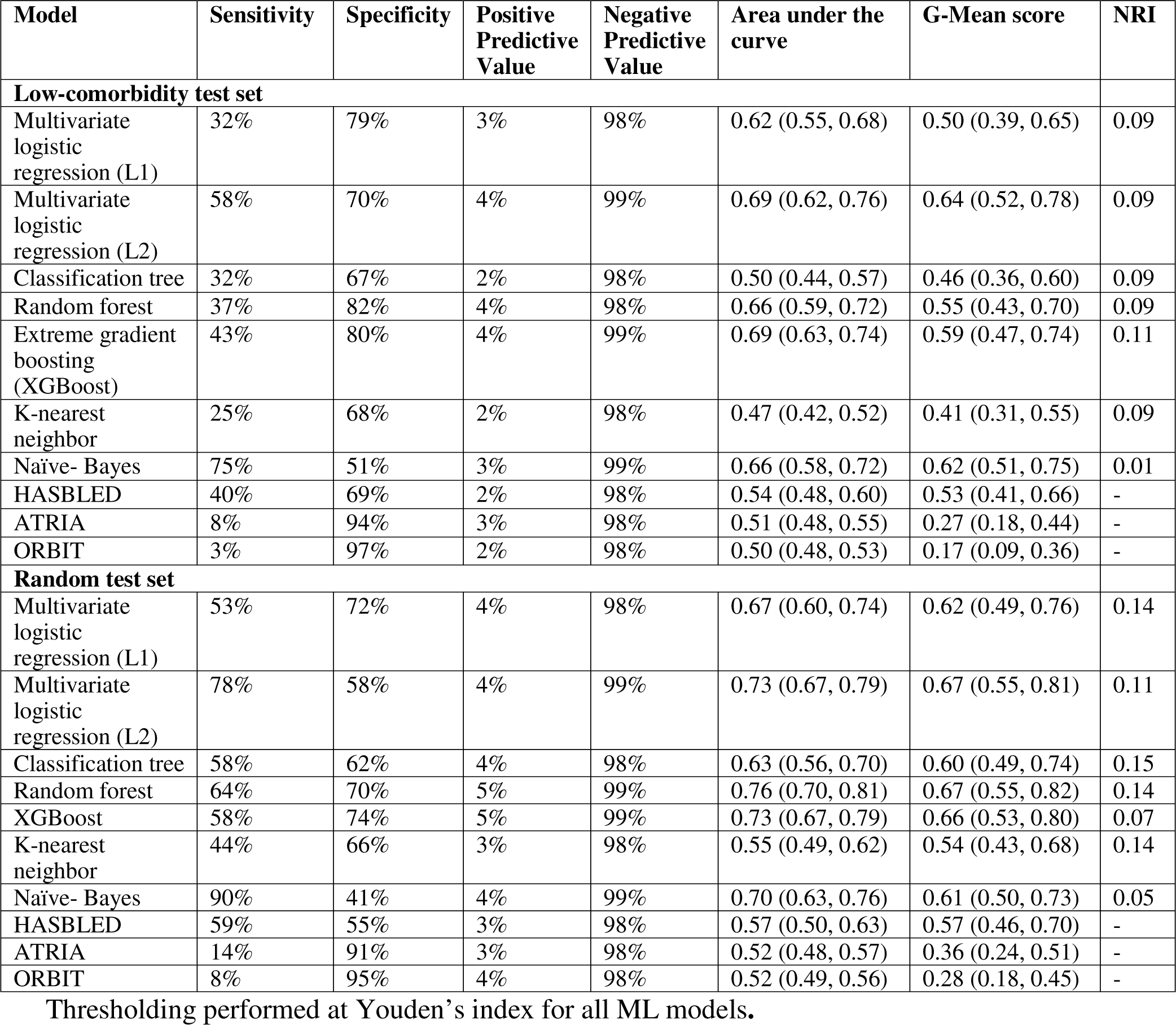
Results of ML models compared with conventional risk scores in predicting bleeding event at 1-year follow-up.

Similarly, in the random cohort, the best performing ML model, random forest (AUC 0.76, 0.70–0.81; G-Mean score 0.67; NRI 0.14; IDI 0.09, Brier score 0.04, log loss 0.21), outperformed the best performing conventional risk score HASBLED (AUC 0.57, 0.50–0.63; G-Mean score 0.57, Brier score 0.45, log loss 16.34), with p-values <0.001 for the individual differences **(Figure 3**, **Table 2, Supplementary Table 3)**.

**Figure 3.**
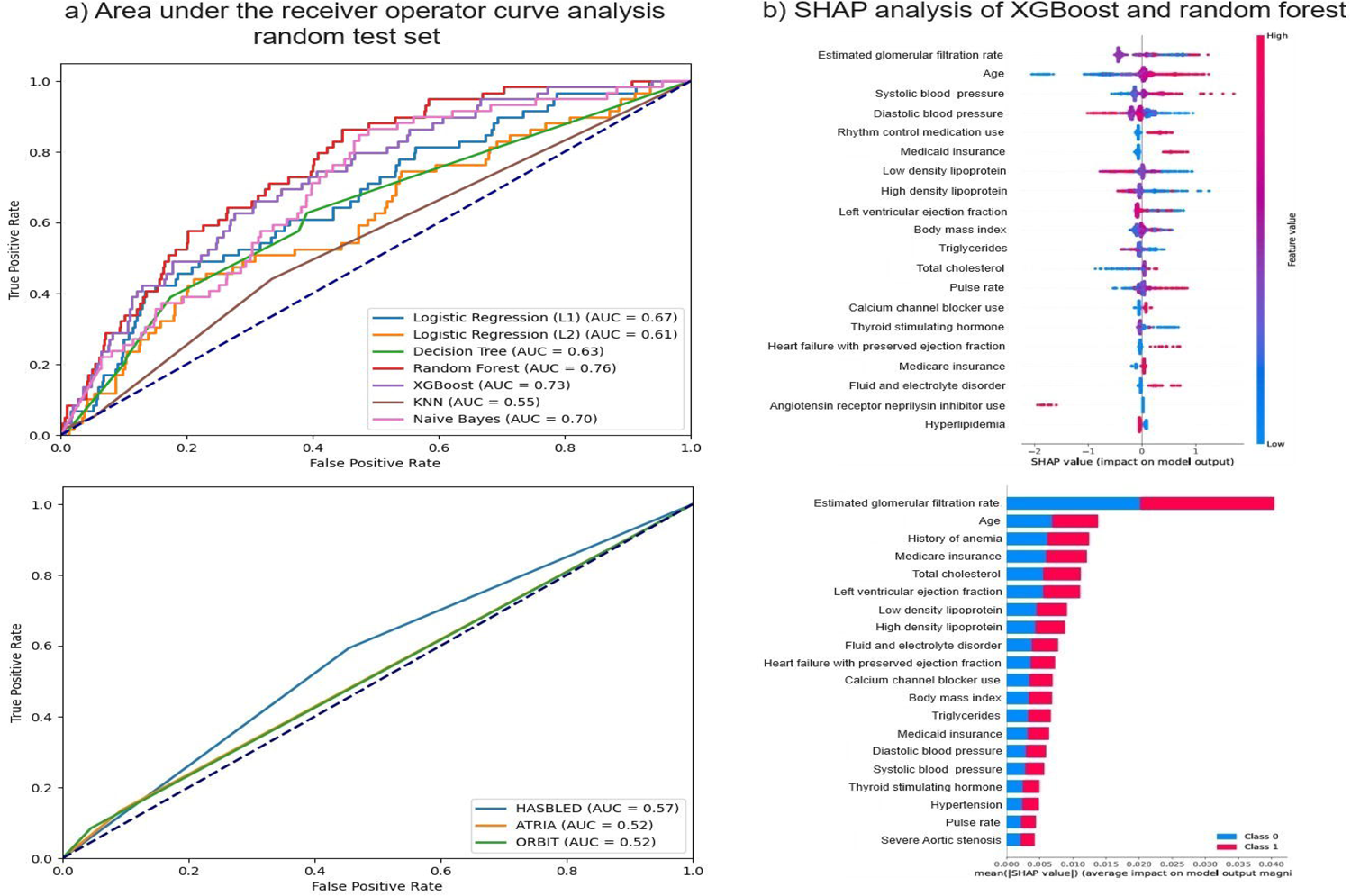
Comparative performance of machine learning models with HASBLED, ORBIT, and ATRIA scores in the random test set to predict a significant bleeding event at 1-year a) AUC-ROC analysis; and b) SHAP analysis for random forest model and extreme gradient boosting (XGBoost) model showing representative factors.

To further assess the performance of the ML models and the HASBLED score, calibration and risk stratification curves were analyzed (**Supplementary Figures 1-3**). The calibration curves demonstrated overall poor calibration in both test cohorts due to underestimation of risk. The predicted probabilities of HASBLED score significantly underestimated the actual risk of bleeding events. In contrast, both XGBoost and random forest models showed improved calibration, with XGBoost displaying the best alignment between predicted and observed probabilities, though some discrepancies were still noted. While all calibration results were suboptimal, the ML models still outperformed the HASBLED score.

Similar results were noted in the risk stratification curves where the overall risk stratification was suboptimal for all models in both cohorts, however, the XGBoost and random forest models demonstrated better alignment between predicted and observed event rates across risk strata compared to HASBLED score. The HASBLED score exhibited substantial underestimation of risk, particularly at the higher end of the risk spectrum, indicating poor risk differentiation and underestimation of high-risk patients.

Other performance metrics including accuracy, F-1 score, AUC-PRC, confusion matrices, IDI, Brier score and log loss for all algorithms are available in **Supplementary Table 3**.

### Secondary Outcomes

The ML models demonstrated superior performance in discriminative power, overall performance, risk stratification, and calibration for bleeding events at 2-year and 5-year follow-up periods including the subgroup with hemorrhagic stroke. At the 2-year follow-up, random forest and XGBoost outperformed conventional risk scores, with an AUC-ROC of 0.70 (95% CI, 0.64-0.76) for XGBoost compared to an AUC-ROC of 0.57 (95% CI, 0.52-0.62) for the best performing conventional HASBLED score. Similar findings were observed at the 5-year follow-up, with random forest and XGBoost as best performing ML models with an AUC-ROC of 0.72 (95% CI, 0.69-0.77) for random forest model compared to 0.57 (95% CI, 0.53-0.61) for HASBLED **(Supplementary Table 4)**.

For hemorrhagic stroke prediction, at the 1-year follow-up, random forest achieved an AUC-ROC of 0.71 (95% CI, 0.60-0.81) while the best-performing conventional score, ORBIT, had an AUC-ROC of 0.58 (95% CI, 0.49-0.71). At the 2-year follow-up, XGBoost outperformed the conventional scores with an AUC-ROC of 0.74 (95% CI, 0.57-0.88) compared to the best-performing conventional score, ATRIA, which had an AUC-ROC of 0.51 (95% CI, 0.47-0.59). Similarly, at the 5-year follow-up, random forest achieved an AUC-ROC of 0.65 (95% CI, 0.53-0.77), while the best-performing conventional score, HASBLED, had an AUC-ROC of 0.56 (95% CI, 0.46-0.66) **(Supplementary Table 5)**.

### Explainability

SHAP analysis was applied to random forest and XGBoost models in both low comorbidity and random test groups to identify the top features in the risk prediction models. The analysis confirmed previously reported risk factors for bleeding, such as older age, renal dysfunction, anemia, hypertension^39–41^. Additionally, novel risk factors were identified, including body mass index, insurance coverage by either Medicare or Medicaid, and dyslipidemia. These factors were identified in both – low-comorbidity and random test sets and by random forest and XGBoost models **(Figure 2 and Figure 3)**. These factors were consistently present at 1-year, 2-year, and 5-year follow-up for bleeding event prediction. Classification tree-schema and logistic regression coefficients for the prediction of 1-year bleeding risk are provided in Supplementary appendix **(Supplementary Figure 3, Supplementary Table 6).** The executable versions of the other machine learning models are not included in this manuscript but are available upon request from the corresponding author.

## Discussion

In this large, real-world cohort study, we demonstrated that ML models, particularly random forest and XGBoost, outperformed conventional risk scores (HAS-BLED, ATRIA, and ORBIT) in predicting clinically significant bleeding events among 24,468 non-valvular AF patients treated with DOACs. The ML models had better discriminative power, overall performance, risk stratification, and calibration compared to the conventional risk scores which was consistent across various follow-up periods (one, two, and five years) and in the subgroup with hemorrhagic stroke. Furthermore, our study identified novel risk factors, such as body mass index, insurance coverage by Medicare or Medicaid, and dyslipidemia, that contributed to improved bleeding risk prediction.

The emergence of alternatives to DOACs, such as left atrial appendage closure devices, underscores the importance of accurate bleeding risk prediction tools to guide personalized treatment decisions ^11,12^. Conventional risk scores, derived from registry data with specific inclusion criteria, often fail to capture the heterogeneity observed in real-world settings. Furthermore, the HAS-BLED score’s reliance on labile INR is outdated, given the shift towards DOACs ^13^. Our findings of a modest improvement in ML performance over conventional risk scores align with prior studies ^16,17^, which consistently demonstrated the poor performance of the HAS-BLED score, with AUC-ROC ranging from 0.50 to 0.64 ^16,19,20,44,45^. However, these studies were limited by their focus on broader contexts or specific AF subpopulations ^18,19^, restricting their applicability to the broader AF population on DOACs in a real-world clinical scenario when first evaluated by a cardiologist for AF management. It is worth noting that in the original HAS-BLED publication, the AUC for the derivation cohort was 0.72, and for the validation cohort, it ranged from 0.50 to 0.67 among patients on warfarin. The performance of the HAS-BLED score in our study, which focused on patients on DOACs, was consistent with the lower end of the validation cohort range from the original publication ^13,46^. Our study addressed these limitations by providing a robust comparison between ML models and multiple conventional risk scores, including HAS-BLED, ATRIA, and ORBIT, using real-world EHR data ^20^. Moreover, we demonstrated the consistent poor performance of conventional risk scores when applied to follow-up periods exceeding one year, which is clinically significant when considering patients for transcatheter left atrial appendage closure ^11,12^.

The superior performance of ML models in capturing complex patterns within real-world clinical data underscores the need for robust, adaptable predictive models that can accommodate individual patient characteristics and evolving therapeutic landscapes. Our diverse cohort, which included patients with varying comorbidities and treatment strategies, emphasizes the potential of ML in enhancing personalized risk assessment and clinical decision-making in managing AF patients. The limitations of conventional risk scores, as revealed by our analysis, highlight the promise of ML models that leverage EHRs to improve predictive accuracy, with ML models demonstrating their ability to employ EHR data effectively, as evidenced by the performance of our models and the identification of novel risk factors through SHAP analysis.

Our study’s pragmatic design utilizes EHRs to develop and evaluate risk prediction models at the patient’s first contact with a Cardiologist. This approach reflects real-world clinical practice where clinicians might not have information on the duration of AF, and patients have already been initiated on DOACs. SHAP analysis identified novel risk factors, such as body mass index, insurance coverage by Medicare or Medicaid, dyslipidemia, and left ventricular ejection fraction on echocardiography, which are not included in conventional risk scores. The variable effect of DOACs in underweight and obese patients has been shown in other studies, prompting guideline recommendations to measure peak and trough levels to ensure that the levels fall in the expected range ^47^. Similarly, factors such as insurance coverage through Medicare and Medicaid and dyslipidemia may indirectly reflect the impact of social determinants of health on the risk of health complications, including bleeding. The consistency of these factors across short and long-term bleeding and hemorrhagic stroke risk in both test cohorts suggests they capture high-risk phenotypes through direct or shared underlying mechanisms.

To ensure the robustness and reliability of our ML models, we employed several strategies to address the challenges inherent in real-world data. To address class imbalance, we utilized different sampling techniques and ratios and performed evaluation with geometric means, enabling a balanced assessment of model performance. We employed 10-fold cross-validation and assessed model performance in test sets with different levels of comorbidities to mitigate overfitting and ensure generalizability. Despite the superior performance of ML models, it is important to acknowledge that the calibration results were suboptimal for both ML models and conventional risk scores. The calibration curves demonstrated an underestimation of risk, particularly for the HAS-BLED score. While the ML models showed improved calibration compared to the HAS-BLED score, there were still discrepancies between the predicted and observed probabilities. Potential reasons for suboptimal calibration include the inherent limitations of real-world data, such as missing or inconsistent data, and the complexity of capturing the dynamic nature of bleeding risk over time. The suboptimal calibration highlights the need for further refinement of the models and emphasizes the importance of interpreting the predictions with caution in clinical practice. Moreover, when implementing the prediction model in clinical practice, poor quality or unavailable input data should be carefully assessed. If key predictors are missing or unreliable, the model’s predictions should be interpreted with caution, and alternative risk assessment methods should be considered. Our study presents a comprehensive array of performance metrics to facilitate an informed discussion of the potential clinical applications of these ML models. By providing an extensive overview of model performance metrics, we aim to support this process and enable a more nuanced understanding of the strengths and limitations of each model in the context of real-world clinical practice.

Building upon the robustness and reliability of our ML models, integrating them into clinical practice presents significant challenges, particularly in selecting the appropriate metric to optimize during the training phase. The choice of metrics depends on the model’s intended application and the utility function it serves, which is shaped by shared decision-making processes between patients and physicians ^48–50^. For example, when predicting major bleeding events, the relative importance of false positives (leading to unnecessary interventions) and false negatives (failing to prevent life-threatening events) may vary based on the patient’s risk profile, comorbidities, and the availability of alternative therapies. These decisions have far-reaching implications, directly influencing treatment choices, resource allocation, and patient well-being. To bridge the gap between theoretical analysis and practical application, engaging patients and physicians in defining the utility function through structured interviews, focus groups, or surveys designed to elicit preferences and values regarding treatment outcomes, side effects, and quality of life is essential. In the absence of a well-defined utility metric, we employed standard metrics to assess the performance of foundational models with minimal hyperparameter fine-tuning, demonstrating that these models outperformed conventional risk estimation approaches. Future elicitation of utility could help develop a mixture of expert models, where individual models optimized for specific metrics are activated according to the elicited utility, realizing the promise of individualized medicine through ML ^51^. The successful integration of ML into clinical decision-making requires a collaborative approach that values the input of all stakeholders and adapts to the ever-evolving landscape of patient care.

## Limitations and Future Directions

Our study has several limitations that should be addressed in future research. It is important to acknowledge that while the ML models outperformed the conventional risk scores, their performance was still limited, with AUC-ROC values ranging from 0.69 to 0.76. These results indicate that there is room for improvement in predicting bleeding risk in AF patients treated with DOACs. The limited performance may be attributed to several factors, such as the complexity of the underlying biological processes, the presence of unknown or unmeasured confounders, and the limitations of the available data. Future studies should focus on refining the models by incorporating additional data sources, exploring novel risk factors, and applying emerging ML techniques to improve predictive accuracy. First, we assumed consistent DOAC therapy throughout the follow-up period, which may not reflect real-world medication adherence and treatment adjustments. To further elucidate the influence of fluctuating DOAC exposure on bleeding risk, subsequent investigations should aim to integrate time-varying covariates into their analyses. Second, while we employed techniques to improve generalizability, relying on a single healthcare system’s EHR data, which included a predominantly white population and low Hispanic ethnicity, may limit the applicability of our findings to other populations. Future studies need external validation to ascertain the robustness and utility of our models in diverse clinical environments. Third, the study population included a mix of patients with varying duration of AF, which may impact their risk profiles. Time series analyses should be conducted in the future to address the changing risk in real-world clinical practice. Fourth, the computation of the HAS-BLED score in our study was limited by the absence of labile INR data, as routine INR monitoring is not recommended for patients on DOACs. This parameter was often missing in real-world practice and was therefore removed from the HAS-BLED score calculation, consistent with prior studies ^52,53^. Similarly, active alcohol use was determined using ICD-9 codes, which may not capture the full spectrum of alcohol consumption. These limitations in computing the HAS-BLED score and alcohol use may have affected the accuracy of these parameters compared to registry data. Fifth, this study did not explicitly address model fairness across different sociodemographic groups. Future research should assess the performance of the models across various subgroups to identify potential biases and explore techniques for mitigating any disparities to ensure equitable predictions for all patients. Finally, while SHAP analysis offers insights into feature importance, improving the explainability of ML models remains an ongoing challenge. Clearer strategies for integrating ML predictions into clinical workflows and decision-making processes are needed, such as developing user-friendly interfaces that present model predictions alongside information obtained from explainability methods such as SHAP.

Future research should also focus on developing a comprehensive utility framework that considers clinical context, patient preferences, and the impact of false positives and false negatives on patient outcomes. This framework will be essential for thresholding and optimizing models for the most relevant metric, ensuring that the ML models are not only accurate but also align with the needs and values of patients and healthcare providers.

In summary, our study demonstrates the superior performance of ML risk models compared to conventional risk scores in predicting clinically significant bleeding events in nonvalvular AF patients treated with DOACs. However, to realize the full potential of ML in clinical practice, future research should focus on addressing limitations such as external validation, model optimization, and improved explainability, while integrating novel data sources, applying emerging ML algorithms, and fostering collaboration among data scientists, clinicians, and patients. Our study lays the foundation for future work to refine these models, define their utility, and translate them into clinical practice to improve patient outcomes and support informed decision-making.

## Data Availability

The data that support the findings of this study are available from the corresponding author upon reasonable request and with permission of the University of Pittsburgh Medical Center.

## Institutional Review Board Statement

The study was conducted following the Declaration of Helsinki and approved by the Institutional Review Board (or Ethics Committee) of University of Pittsburgh.

## Informed Consent Statement

Not applicable

## Acknowledgments

Not applicable.

## Protocol

The study protocol is available upon request from the corresponding author.

## Registration

The study was not registered.

## Code sharing

The analytical code is available from the corresponding author upon reasonable request.

