## Supplementary appendix for "Machine Learning - Based Bleeding Risk Predictions in Atrial Fibrillation Patients on Direct Oral Anticoagulants"

**Supplementary Tables**

- **Supplementary Table 1.** ICD-9-CM and ICD-10-CM codes for different study outcomes.
- **Supplementary Table 2.** Variables with percent missing values, other variables had no missing values.
- **Supplementary Table 3.** Results of ML models compared with conventional risk scores in predicting bleeding event at 1-year follow-up.
- **Supplementary Table 4.** Results of ML models compared with conventional risk scores in predicting bleeding event at 2-years and 5-years of follow-up.
- **Supplementary Table 5.** Results of ML models compared with conventional risk scores in predicting hemorrhagic stroke at 1-year, 2-year and 5-years of follow-up.
- **Supplementary Table 6.** Equations for the multivariate logistic regression equations for the final models.

**Supplementary Figures**

- **Supplementary Figure 1.** Calibration plots and risk stratification plots for extreme gradient boosting (XGBoost) in the low comorbid and random test cohorts for prediction of 1-year bleeding risk.
- **Supplementary Figure 2.** Calibration plots and risk stratification plots for random forest model in the low comorbid and random test cohorts for prediction of 1-year bleeding risk.
- **Supplementary Figure 3.** Classification tree schema for prediction of major bleeding event at 1-year.

**Supplementary Table 1.** ICD-9-CM and ICD-10-CM codes for different study outcomes.

**Definition of study outcomes**

The study outcomes (apart from all-cause mortality) were defined using the 9^th^ and 10^th^ revisions of the International Classification of Diseases, Clinical Modification (ICD-9-CM and ICD-10-CM) as detailed in Supplemental Table 1 below.

| **Outcome** | **ICD-9-CM** | **ICD-10-CM** |
| --- | --- | --- |
| **Significant Bleeding Events*** | 336.1, 423, 430, 432, 432.1, 432.9, 456, 456.2, 530.7, 530.82, 531.0, 531.00, 531.01, 531.1, 531.10, 531.11, 531.2, 531.20, 531.21, 531.3, 531.30, 531.31, 531.4, 531.40, 531.41, 531.5, 531.50, 531.51, 531.6, 531.60, 531.61, 532.0, 532.00, 532.01, 532.1, 532.10, 532.11, 532.2, 532.20, 532.21, 532.3, 532.30, 532.31, 532.4, 532.40, 532.41, 532.5, 532.50, 532.51, 532.6, 532.60, 532.61, 533.0, 533.00, 533.01, 533.1, 533.10, 533.11, 533.2, 533.20, 533.21, 533.3, 533.30, 533.31, 533.4, 533.40, 533.41, 533.5, 533.50, 533.51, 533.6, 533.60, 533.61, 534.0, 534.00, 534.01, 534.1, 534.10, 534.11, 534.2, 534.20, 534.21, 534.3, 534.30, 534.31, 534.4, 534.40, 534.41, 534.5, 534.51, 534.6, 534.60, 534.61, 535.0, 535.00, 535.01, 535.1, 535.10, 535.11, 535.2, 535.20, 535.21, 535.3, 535.30, 535.31, 535.4, 535.40, 535.41, 535.5, 535.50, 535.51, 535.6, 535.60, 535.61, 537.84, 562.02, 562.03, 562.12, 562.13, 568.81, 569.3, 569.85, 578, 578.1, 578.9, 767.11, 852, 852.01, 852.02, 852.03, 852.04, 852.05, 852.06, 852.09, 852.1, 852.11, 852.12, 852.13, 852.14, 852.15, 852.16, 852.19, 852.2, 852.21, 852.22, 852.23, 852.24, 852.25, 852.26, 852.29, 852.3, 852.31, 852.32, 852.33, 852.34, 852.35, 852.36, 852.39, 852.4, 852.41, 852.42, 852.43, 852.44, 852.45, 852.46, 852.49, 852.5, 852.51, 852.52, 852.53, 852.54, 852.55, 852.56, 852.59, 853, 853.01, 853.02, 853.03, 853.04, 853.05, 853.06, 853.09, 853.1, 853.11, 853.12, 853.13, 853.14, 853.15, 853.16, 853.19, 996.72, 996.74, 998.11 | G95.19, I31.2, I60.8, I60.9, I62, I62.0, I62.00, I62.01, I62.02, I62.03, I62.1, I62.9, I85.01, I97.618, K22.6, K22.8, K25.0, K25.1, K25.2, K25.3, K25.4, K25.5, K25.6, K26.0, K26.2, K26.3, K26.4, K26.5, K26.6, K27.0, K27.1, K27.2, K27.3, K27.4, K27.5, K27.6, K28.0, K28.1, K28.2, K28.3, K28.4, K28.5, K28.9, K29.00, K29.01, K29.30, K29.31, K29.40, K29.41, K29.50, K29.51, K29.60, K29.61, K29.70, K29.71, K29.80, K29.81, K29.90, K29.91, K31.811, K31.82, K55.21, K57.01, K57.11, K57.13, K57.21, K57.31, K57.33, K57.41, K57.51, K57.53, K57.91, K57.93, K62.5, K66.1, K92.1, K92.2, P12.2, R58, S06.340A, S06.341A, S06.342A, S06.343A, S06.344A, S06.345A, S06.346A, S06.347A, S06.348A, S06.349A, S06.350A, S06.351A, S06.352A, S06.353A, S06.354A, S06.355A, S06.356A, S06.357A, S06.358A, S06.359A, S06.360A, S06.361A, S06.362A, S06.363A, S06.364A, S06.365A, S06.366A, S06.367A, S06.368A, S06.369A, S06.370A, S06.371A, S06.372A, S06.373A, S06.374A, S06.375A, S06.376A, S06.377A, S06.378A, S06.379A, S06.380A, S06.381A, S06.382A, S06.383A, S06.384A, S06.385A, S06.386A, S06.387A, S06.388A, S06.389A, S06.4X0A, S06.4X1A, S06.4X2A, S06.4X3A, S06.4X4A, S06.4X5A, S06.4X6A, S06.4X7A, S06.4X8A, S06.4X9A, S06.5X0A, S06.5X1A, S06.5X2A, S06.5X3A, S06.5X4A, S06.5X5A, S06.5X6A, S06.5X7A, S06.5X8A, S06.5X9A, S06.6X0A, S06.6X1A, S06.6X2A, S06.6X3A, S06.6X4A, S06.6X5A, S06.6X6A, S06.6X7A, S06.6X8A, S06.6X9A, T82.837A, T82.838A |
| **Hemorrhagic Stroke** | 430, 431, 432.0, 432.1, 432.9, 444.01, 444.09, 444.1, 444.21, 444.22, 444.81, 444.89, 444.9 | I60.00, I60.01, I60.02, I60.10, I60.11, I60.12, I60.2, I60.30, I60.31, I60.32, I60.4, I60.50, I60.51, I60.52, I60.6, I60.7, I60.8, I60.9, I61.0, I61.1, I61.2, I61.3, I61.4, I61.5, I61.6, I61.8, I61.9, I62.00, I62.01, I62.02, I62.03, I62.1, I62.9 |

*Bleeding events were considered significant if one of the corresponding ICD codes was listed as one of the top 3 diagnoses during an inpatient admission

ICD = International Classification of Diseases

**Supplementary Table 2.** Variables with percent missing values, other variables had no missing values.

| **Variable** | **Percent Missing** |
| --- | --- |
| Transthoracic echocardiography |  |
| Moderate to Severe Mitral Stenosis | 99.3 |
| Left Atrial Diameter (millimeters) | 75.0 |
| Left Ventricular Systolic Diameter (millimeters) | 62.9 |
| Left Ventricular Diastolic Diameter (millimeters) | 62.7 |
| Moderate to Severe Mitral Regurgitation | 60.7 |
| Left Ventricular Ejection Fraction (%) | 54.3 |
| Hemoglobin A1C | 60.6 |
| Thyroid Stimulating Hormone (micro international units per milliliter) | 45.5 |
| Lipid panel (mg/dl) |  |
| Low-Density Lipoprotein Cholesterol | 44.3 |
| High-Density Lipoprotein Cholesterol | 44.2 |
| Triglycerides | 43.8 |
| Total Cholesterol | 43.4 |
| Estimated Glomerular Filtration Rate (milliliters per minute per 1.73 square meters) | 37.2 |
| Illicit Drug Use | 14.4 |
| Heart rate (beats per minute) | 9.5 |
| Heart Failure history | 8.7 |
| Alcohol Use | 8.1 |
| Hispanic or Latino Ethnic Group | 3.5 |
| Smoker | 2.0 |
| Body Mass Index (kilograms per square meter) | 1.9 |
| Blood pressure (systolic and diastolic) | 1.1 |
| Race | 0.7 |

**Supplementary Table 3.** Results of ML models compared with conventional risk scores in predicting bleeding event at 1-year follow-up.

| **Model** | **Accuracy** | **F-1 score** | **AUC-PRC** | **Confusion matrix (TP, FP, FN, TN)** | **IDI** | **Brier score** | **Log loss** |
| --- | --- | --- | --- | --- | --- | --- | --- |
| **Low-comorbidity test set** | | | | | | | |
| Multivariate logistic regression (L1) | 78% | 0.05 | 0.03 | [[2375, 619], [41, 19]] | 0.07 | 0.04 | 0.20 |
| Multivariate logistic regression (L2) | 69% | 0.07 | 0.04 | [[2086, 908], [25, 35]] | 0.04 | 0.04 | 0.19 |
| Classification tree | 66% | 0.04 | 0.02 | [[2011, 983], [41, 19]] | 0.08 | 0.04 | 0.20 |
| Random forest | 82% | 0.07 | 0.04 | [[2469, 525], [38, 22]] | 0.06 | 0.04 | 0.19 |
| Extreme gradient boosting (XGBoost) | 79% | 0.07 | 0.03 | [[2383, 611], [34, 26]] | 0.04 | 0.04 | 0.19 |
| K-nearest neighbor | 67% | 0.03 | 0.02 | [[2043, 951], [45, 15]] | 0.11 | 0.07 | 0.80 |
| Naïve- Bayes | 52% | 0.06 | 0.03 | [[1528, 1466], [15, 45]] | 0.01 | 0.78 | 6.51 |
| HASBLED | 68% | 0.05 | 0.02 | [[2058, 936], [36, 24]] | - | 0.32 | 11.47 |
| ATRIA | 92% | 0.04 | 0.02 | [[2817, 177], [55, 5]] | - | 0.05 | 1.72 |
| ORBIT | 95% | 0.03 | 0.02 | [[2906, 88], [58, 2]] | - | 0.08 | 2.74 |
| **Random test set** | | | | | | | |
| Multivariate logistic regression (L1) | 71% | 0.08 | 0.05 | [[1767, 696], [28, 31]] | 0.10 | 0.05 | 0.22 |
| Multivariate logistic regression (L2) | 58% | 0.08 | 0.06 | [[1427, 1036], [13, 46]] | 0.07 | 0.05 | 0.22 |
| Classification tree | 62% | 0.07 | 0.04 | [[1532, 931], [25, 34]] | 0.09 | 0.05 | 0.23 |
| Random forest | 70% | 0.09 | 0.07 | [[1722, 741], [21, 38]] | 0.09 | 0.04 | 0.21 |
| XGBoost | 73% | 0.09 | 0.06 | [[1818, 645], [25, 34]] | 0.03 | 0.05 | 0.21 |
| K-nearest neighbor | 66% | 0.06 | 0.03 | [[1637, 826], [33, 26]] | 0.10 | 0.07 | 0.79 |
| Naïve- Bayes | 42% | 0.07 | 0.05 | [[1002, 1461], [6, 53]] | 0.02 | 0.80 | 8.63 |
| HASBLED | 55% | 0.06 | 0.03 | [[1344, 1119], [24, 35]] | - | 0.45 | 16.34 |
| ATRIA | 89% | 0.05 | 0.02 | [[2233, 230], [51, 8]] | - | 0.07 | 2.36 |
| ORBIT | 93% | 0.06 | 0.03 | [[2352, 111], [54, 5]] | - | 0.11 | 4.02 |

Thresholding at Youden’s index for all ML models**.**

**Supplementary Table 4.** Results of ML models compared with conventional risk scores in predicting bleeding event at 2-years and 5-years of follow-up.

| **Model** | **Time (yrs)** | **AUC** | **Sensitivity** | **Specificity** | **PPV** | **NPV** | **G-Mean score** | | **NRI** | | **Brier score** |
| --- | --- | --- | --- | --- | --- | --- | --- | --- | --- | --- | --- |
| **Low-comorbidity test set** | | | | | | | | | | | |
| Multivariate logistic regression (L1) | 2 | 0.50 (0.44, 0.57) | 24% | 78% | 3% | 97% | 0.43 (0.34, 0.54) | | 0.14 | | 0.05 |
| Multivariate logistic regression (L1) | 5 | 0.54 (0.5, 0.59) | 21% | 83% | 7% | 95% | 0.42 (0.35, 0.51) | | 0.15 | | 0.07 |
| Multivariate logistic regression (L2) | 2 | 0.65 (0.59, 0.7) | 59% | 68% | 6% | 98% | 0.63 (0.54, 0.75) | | 0.14 | | 0.05 |
| Multivariate logistic regression (L2) | 5 | 0.69 (0.65, 0.73) | 21% | 92% | 14% | 95% | 0.44 (0.36, 0.53) | | 0.15 | | 0.06 |
| Classification tree | 2 | 0.67 (0.62, 0.73) | 64% | 65% | 6% | 98% | 0.64 (0.55, 0.76) | | 0.14 | | 0.05 |
| Classification tree | 5 | 0.68 (0.64, 0.72) | 55% | 70% | 10% | 96% | 0.62 (0.55, 0.71) | | 0.13 | | 0.06 |
| Random forest | 2 | 0.69 (0.63, 0.74) | 47% | 82% | 8% | 98% | 0.62 (0.52, 0.74) | | 0.14 | | 0.04 |
| Random forest | 5 | 0.71 (0.67, 0.76) | 52% | 78% | 12% | 97% | 0.64 (0.56, 0.73) | | 0.15 | | 0.06 |
| Extreme gradient boosting (XGBoost) | 2 | 0.70 (0.64, 0.76) | 47% | 82% | 8% | 98% | 0.62 (0.52, 0.74) | | 0.10 | | 0.05 |
| XGBoost | 5 | 0.72 (0.68, 0.76) | 53% | 77% | 12% | 97% | 0.64 (0.56, 0.73) | | 0.11 | | 0.06 |
| K-nearest neighbor (KNN) | 2 | 0.53 (0.49, 0.58) | 39% | 68% | 4% | 97% | 0.51 (0.43, 0.62) | | 0.15 | | 0.08 |
| KNN | 5 | 0.55 (0.51, 0.59) | 44% | 64% | 7% | 95% | 0.53 (0.46, 0.61) | | 0.09 | | 0.10 |
| Naïve- Bayes | 2 | 0.60 (0.54, 0.66) | 57% | 60% | 4% | 98% | 0.58 (0.49, 0.69) | | 0.01 | | 0.56 |
| Naïve- Bayes | 5 | 0.65 (0.61, 0.7) | 58% | 67% | 9% | 96% | 0.62 (0.55, 0.71) | | 0.03 | | 0.17 |
| HASBLED | 2 | 0.57 (0.52, 0.62) | 45% | 69% | 4% | 97% | 0.56 (0.46, 0.67) | | - | | 0.31 |
| HASBLED | 5 | 0.57 (0.53, 0.61) | 45% | 69% | 8% | 96% | 0.56 (0.49, 0.64) | | - | | 0.32 |
| ATRIA | 2 | 0.53 (0.50, 0.57) | 12% | 95% | 7% | 97% | 0.34 (0.25, 0.46) | | - | | 0.05 |
| ATRIA | 5 | 0.54 (0.51, 0.56) | 12% | 96% | 14% | 95% | 0.34 (0.27, 0.43) | | - | | 0.07 |
| ORBIT | 2 | 0.51 (0.49, 0.54) | 5% | 98% | 7% | 97% | 0.22 (0.15, 0.36) | | - | | 0.08 |
| ORBIT | 5 | 0.51 (0.5, 0.53) | 5% | 98% | 12% | 95% | 0.22 (0.16, 0.32) | | - | | 0.09 |
| **Random test set** | | | | | | | |  | |  | |
| Multivariate logistic regression (L1) | 2 | 0.52 (0.46, 0.59) | 42% | 62% | 4% | 97% | 0.51 (0.42, 0.62) | | 0.13 | | 0.06 |
| Multivariate logistic regression (L1) | 5 | 0.54 (0.49, 0.59) | 29% | 78% | 7% | 95% | 0.48 (0.4, 0.57) | | 0.13 | | 0.07 |
| Multivariate logistic regression (L2) | 2 | 0.65 (0.59, 0.72) | 70% | 52% | 5% | 98% | 0.60 (0.52, 0.71) | | 0.13 | | 0.06 |
| Multivariate logistic regression (L2) | 5 | 0.65 (0.61, 0.7) | 35% | 81% | 10% | 95% | 0.53 (0.45, 0.63) | | 0.13 | | 0.07 |
| Classification tree | 2 | 0.66 (0.6, 0.72) | 68% | 58% | 6% | 98% | 0.63 (0.54, 0.74) | | 0.10 | | 0.06 |
| Classification tree | 5 | 0.66 (0.61, 0.7) | 55% | 65% | 9% | 96% | 0.60 (0.52, 0.69) | | 0.10 | | 0.07 |
| Random forest | 2 | 0.69 (0.62, 0.74) | 57% | 70% | 7% | 98% | 0.63 (0.53, 0.75) | | 0.13 | | 0.05 |
| Random forest | 5 | 0.69 (0.65, 0.73) | 53% | 69% | 9% | 96% | 0.60 (0.52, 0.7) | | 0.13 | | 0.06 |
| Extreme gradient boosting (XGBoost) | 2 | 0.68 (0.62, 0.73) | 58% | 72% | 7% | 98% | 0.65 (0.55, 0.77) | | 0.07 | | 0.06 |
| XGBoost | 5 | 0.70 (0.66, 0.74) | 58% | 69% | 10% | 96% | 0.63 (0.55, 0.73) | | 0.08 | | 0.07 |
| K-nearest neighbor(KNN) | 2 | 0.58 (0.52, 0.64) | 50% | 64% | 5% | 97% | 0.57 (0.47, 0.68) | | 0.05 | | 0.09 |
| KNN | 5 | 0.54 (0.49, 0.59) | 45% | 62% | 7% | 95% | 0.53 (0.45, 0.62) | | 0.09 | | 0.10 |
| Naïve- Bayes | 2 | 0.62 (0.56, 0.68) | 72% | 47% | 5% | 98% | 0.58 (0.49, 0.68) | | 0.01 | | 0.64 |
| Naïve- Bayes | 5 | 0.64 (0.59, 0.68) | 67% | 54% | 8% | 96% | 0.60 (0.53, 0.69) | | 0.01 | | 0.26 |
| HASBLED | 2 | 0.56 (0.52, 0.62) | 56% | 57% | 5% | 97% | 0.56 (0.47, 0.67) | | - | | 0.43 |
| HASBLED | 5 | 0.56 (0.52, 0.61) | 55% | 57% | 7% | 96% | 0.56 (0.49, 0.65) | | - | | 0.43 |
| ATRIA | 2 | 0.57 (0.53, 0.62) | 24% | 91% | 9% | 97% | 0.47 (0.37, 0.59) | | - | | 0.07 |
| ATRIA | 5 | 0.53 (0.5, 0.56) | 15% | 92% | 9% | 95% | 0.37 (0.29, 0.46) | | - | | 0.09 |
| ORBIT | 2 | 0.52 (0.49, 0.55) | 8% | 96% | 7% | 97% | 0.28 (0.19, 0.41) | | - | | 0.11 |
| ORBIT | 5 | 0.50 (0.48, 0.52) | 4% | 96% | 6% | 94% | 0.20 (0.12, 0.3) | | - | | 0.13 |

Thresholding at Youden’s index for all ML models**.**

**Supplementary Table 5.** Results of ML models compared with conventional risk scores in predicting hemorrhagic stroke at 1-year, 2-year and 5-years of follow-up.

| **Model** | **Time (yrs)** | **AUC** | **Sensitivity** | **Specificity** | **PPV** | **NPV** | **G-Mean score** | **NRI** | **Brier score** |
| --- | --- | --- | --- | --- | --- | --- | --- | --- | --- |
| **Low-comorbidity test set** | | | | | | | | | |
| Multivariate logistic regression (L1) | 1 | 0.56 (0.38, 0.72) | 27% | 76% | 0% | 100% | 0.45 (0.24, 0.86) | 0.06 | 0.03 |
| Multivariate logistic regression (L1) | 2 | 0.5 (0.38, 0.63) | 21% | 75% | 0% | 100% | 0.4 (0.21, 0.75) | 0.01 | 0.06 |
| Multivariate logistic regression (L1) | 5 | 0.56 (0.45, 0.67) | 33% | 81% | 1% | 99% | 0.52 (0.35, 0.78) | 0.12 | 0.05 |
| Multivariate logistic regression (L2) | 1 | 0.64 (0.46, 0.83) | 0% | 100% | 0% | 100% | - | 0.06 | 0.03 |
| Multivariate logistic regression (L2) | 2 | 0.63 (0.46, 0.79) | 0% | 100% | 0% | 100% | - | 0.01 | 0.03 |
| Multivariate logistic regression (L2) | 5 | 0.58 (0.45, 0.7) | 29% | 86% | 2% | 99% | 0.5 (0.33, 0.76) | 0.12 | 0.03 |
| Classification tree | 1 | 0.67 (0.51, 0.83) | 64% | 67% | 1% | 100% | 0.65 (0.41, 1.04) | 0.10 | 0.04 |
| Classification tree | 2 | 0.55 (0.42, 0.7) | 36% | 72% | 1% | 100% | 0.51 (0.3, 0.84) | 0.06 | 0.04 |
| Classification tree | 5 | 0.61 (0.49, 0.72) | 50% | 73% | 2% | 99% | 0.6 (0.43, 0.85) | 0.12 | 0.04 |
| Random forest | 1 | 0.71 (0.6, 0.81) | 18% | 86% | 0% | 100% | 0.39 (0.19, 0.84) | 0.06 | 0.03 |
| Random forest | 2 | 0.66 (0.51, 0.81) | 29% | 82% | 1% | 100% | 0.49 (0.28, 0.84) | 0.01 | 0.03 |
| Random forest | 5 | 0.65 (0.53, 0.77) | 42% | 78% | 2% | 99% | 0.57 (0.4, 0.83) | 0.12 | 0.03 |
| Extreme gradient boosting (XGBoost) | 1 | 0.54 (0.36, 0.72) | 18% | 90% | 1% | 100% | 0.4 (0.19, 0.86) | 0.09 | 0.04 |
| XGBoost | 2 | 0.74 (0.57, 0.88) | 36% | 88% | 1% | 100% | 0.56 (0.34, 0.93) | 0.13 | 0.03 |
| XGBoost | 5 | 0.62 (0.5, 0.73) | 38% | 76% | 1% | 99% | 0.54 (0.36, 0.78) | 0.13 | 0.04 |
| K-nearest neighbor(KNN) | 1 | 0.51 (0.37, 0.67) | 36% | 64% | 0% | 100% | 0.48 (0.27, 0.85) | 0.03 | 0.06 |
| KNN | 2 | 0.68 (0.55, 0.8) | 86% | 47% | 1% | 100% | 0.64 (0.43, 0.94) | 0.02 | 0.05 |
| KNN | 5 | 0.5 (0.4, 0.61) | 33% | 64% | 1% | 99% | 0.46 (0.31, 0.69) | 0.05 | 0.07 |
| Naïve- Bayes | 1 | 0.63 (0.42, 0.83) | 27% | 84% | 1% | 100% | 0.48 (0.25, 0.91) | 0.15 | 0.90 |
| Naïve- Bayes | 2 | 0.57 (0.4, 0.74) | 29% | 86% | 1% | 100% | 0.5 (0.28, 0.86) | 0.10 | 0.91 |
| Naïve- Bayes | 5 | 0.57 (0.45, 0.7) | 21% | 90% | 2% | 99% | 0.43 (0.27, 0.7) | 0.07 | 0.94 |
| HASBLED | 1 | 0.53 (0.35, 0.69) | 36% | 70% | 0% | 100% | 0.5 (0.28, 0.89) | - | 0.30 |
| HASBLED | 2 | 0.49 (0.39, 0.63) | 29% | 70% | 0% | 100% | 0.45 (0.26, 0.78) | - | 0.30 |
| HASBLED | 5 | 0.56 (0.46, 0.66) | 42% | 70% | 1% | 99% | 0.54 (0.37, 0.78) | - | 0.30 |
| ATRIA | 1 | 0.57 (0.47, 0.7) | 18% | 95% | 1% | 100% | 0.41 (0.2, 0.88) | - | 0.03 |
| ATRIA | 2 | 0.51 (0.47, 0.59) | 7% | 95% | 1% | 100% | 0.26 (0.09, 0.72) | - | 0.03 |
| ATRIA | 5 | 0.5 (0.47, 0.55) | 4% | 95% | 1% | 99% | 0.19 (0.07, 0.54) | - | 0.06 |
| ORBIT | 1 | 0.58 (0.49, 0.71) | 18% | 97% | 2% | 100% | 0.42 (0.2, 0.89) | - | 0.05 |
| ORBIT | 2 | 0.49 (0.49, 0.49) | 0% | 100% | 0% | 100% | - | - | 0.05 |
| ORBIT | 5 | 0.51 (0.49, 0.56) | 4% | 98% | 1% | 99% | 0.2 (0.07, 0.55) | - | 0.03 |
| **Random test set** | | | | | | | | | |
| Multivariate logistic regression (L1) | 1 | 0.59 (0.47, 0.7) | 42% | 72% | 1% | 99% | 0.55 (0.36, 0.83) | 0.11 | 0.04 |
| Multivariate logistic regression (L1) | 2 | 0.56 (0.46, 0.66) | 78% | 29% | 1% | 99% | 0.48 (0.36, 0.63) | 0.27 | 0.06 |
| Multivariate logistic regression (L1) | 5 | 0.54 (0.46, 0.62) | 33% | 78% | 3% | 98% | 0.51 (0.38, 0.67) | 0.10 | 0.06 |
| Multivariate logistic regression (L2) | 1 | 0.62 (0.49, 0.74) | 0% | 100% | 0% | 99% | - | 0.11 | 0.04 |
| Multivariate logistic regression (L2) | 2 | 0.68 (0.58, 0.77) | 0% | 100% | 0% | 99% | - | 0.27 | 0.04 |
| Multivariate logistic regression (L2) | 5 | 0.64 (0.57, 0.71) | 43% | 73% | 3% | 98% | 0.56 (0.43, 0.72) | 0.10 | 0.04 |
| Classification tree | 1 | 0.59 (0.47, 0.71) | 84% | 26% | 1% | 100% | 0.47 (0.34, 0.65) | 0.13 | 0.06 |
| Classification tree | 2 | 0.41 (0.32, 0.53) | 22% | 66% | 1% | 99% | 0.38 (0.25, 0.6) | 0.23 | 0.05 |
| Classification tree | 5 | 0.6 (0.52, 0.67) | 45% | 70% | 3% | 98% | 0.56 (0.44, 0.72) | 0.10 | 0.05 |
| Random forest | 1 | 0.64 (0.5, 0.77) | 42% | 77% | 1% | 99% | 0.57 (0.38, 0.86) | 0.11 | 0.04 |
| Random forest | 2 | 0.64 (0.54, 0.75) | 41% | 72% | 2% | 99% | 0.54 (0.38, 0.77) | 0.27 | 0.04 |
| Random forest | 5 | 0.65 (0.58, 0.73) | 55% | 67% | 4% | 99% | 0.61 (0.48, 0.77) | 0.10 | 0.04 |
| Extreme gradient boosting (XGBoost) | 1 | 0.64 (0.52, 0.76) | 21% | 88% | 1% | 99% | 0.43 (0.25, 0.74) | 0.07 | 0.04 |
| XGBoost | 2 | 0.66 (0.56, 0.76) | 33% | 83% | 2% | 99% | 0.52 (0.36, 0.77) | 0.15 | 0.05 |
| XGBoost | 5 | 0.65 (0.57, 0.73) | 41% | 77% | 4% | 98% | 0.56 (0.43, 0.73) | 0.09 | 0.04 |
| K-nearest neighbor (KNN) | 1 | 0.59 (0.46, 0.7) | 53% | 64% | 1% | 99% | 0.58 (0.4, 0.85) | 0.15 | 0.07 |
| KNN | 2 | 0.46 (0.36, 0.58) | 44% | 46% | 1% | 99% | 0.45 (0.32, 0.63) | 0.26 | 0.06 |
| KNN | 5 | 0.52 (0.45, 0.6) | 45% | 60% | 2% | 98% | 0.52 (0.4, 0.66) | 0.09 | 0.09 |
| Naïve- Bayes | 1 | 0.56 (0.43, 0.68) | 37% | 73% | 1% | 99% | 0.52 (0.34, 0.8) | 0.14 | 0.86 |
| Naïve- Bayes | 2 | 0.67 (0.57, 0.75) | 41% | 76% | 2% | 99% | 0.56 (0.39, 0.79) | 0.20 | 0.88 |
| Naïve- Bayes | 5 | 0.57 (0.48, 0.65) | 24% | 83% | 3% | 98% | 0.45 (0.33, 0.62) | 0.10 | 0.90 |
| HASBLED | 1 | 0.45 (0.34, 0.56) | 32% | 58% | 1% | 99% | 0.43 (0.27, 0.68) | - | 0.42 |
| HASBLED | 2 | 0.63 (0.54, 0.71) | 70% | 57% | 2% | 99% | 0.63 (0.47, 0.85) | - | 0.43 |
| HASBLED | 5 | 0.55 (0.47, 0.62) | 53% | 57% | 3% | 98% | 0.55 (0.43, 0.69) | - | 0.43 |
| ATRIA | 1 | 0.56 (0.47, 0.67) | 21% | 90% | 2% | 99% | 0.43 (0.25, 0.75) | - | 0.05 |
| ATRIA | 2 | 0.51 (0.45, 0.58) | 11% | 91% | 1% | 99% | 0.32 (0.17, 0.58) | - | 0.05 |
| ATRIA | 5 | 0.51 (0.47, 0.55) | 10% | 91% | 2% | 98% | 0.3 (0.19, 0.48) | - | 0.11 |
| ORBIT | 1 | 0.5 (0.47, 0.56) | 5% | 95% | 1% | 99% | 0.22 (0.08, 0.61) | - | 0.10 |
| ORBIT | 2 | 0.52 (0.48, 0.57) | 7% | 96% | 2% | 99% | 0.26 (0.13, 0.55) | - | 0.10 |
| ORBIT | 5 | 0.5 (0.48, 0.53) | 4% | 96% | 2% | 98% | 0.2 (0.1, 0.4) | - | 0.06 |

Thresholding at Youden’s index for all ML models**.**

**Supplementary Table 6.** Equations for the multivariate logistic regression equations for the final models.

| **Model** | **Variable** | **Coefficient** |
| --- | --- | --- |
| Multivariate logistic regression (L1) | Age | 0.014 |
|  | Blood pressure (Systolic) | 0.006 |
|  | Cholesterol level | -0.005 |
|  | LV ejection fraction | -0.008 |
|  | EGFR | -0.014 |
|  | Blood pressure (Diastolic) | -0.019 |
| Multivariate logistic regression (L2) | Medicaid insurance | 0.150 |
|  | Age | 0.140 |
|  | Heart failure with preserved ejection fraction | 0.125 |
|  | Medicare insurance | 0.123 |
|  | Anemia history | 0.110 |
|  | Blood pressure (Systolic) | 0.100 |
|  | History of severe aortic stenosis | 0.094 |
|  | Active cancer | 0.083 |
|  | Calcium channel blocker | 0.080 |
|  | Prediabetes | 0.079 |
|  | Hypertension | 0.074 |
|  | Insulin use | 0.064 |
|  | EGFR | -0.083 |
|  | Blood pressure (Diastolic) | -0.095 |
|  | ARNI | -0.128 |

**Supplementary Figure 1.** Calibration plots and risk stratification plots for extreme gradient boosting (XGBoost) in the low comorbid and random test cohorts for prediction of 1-year bleeding risk.

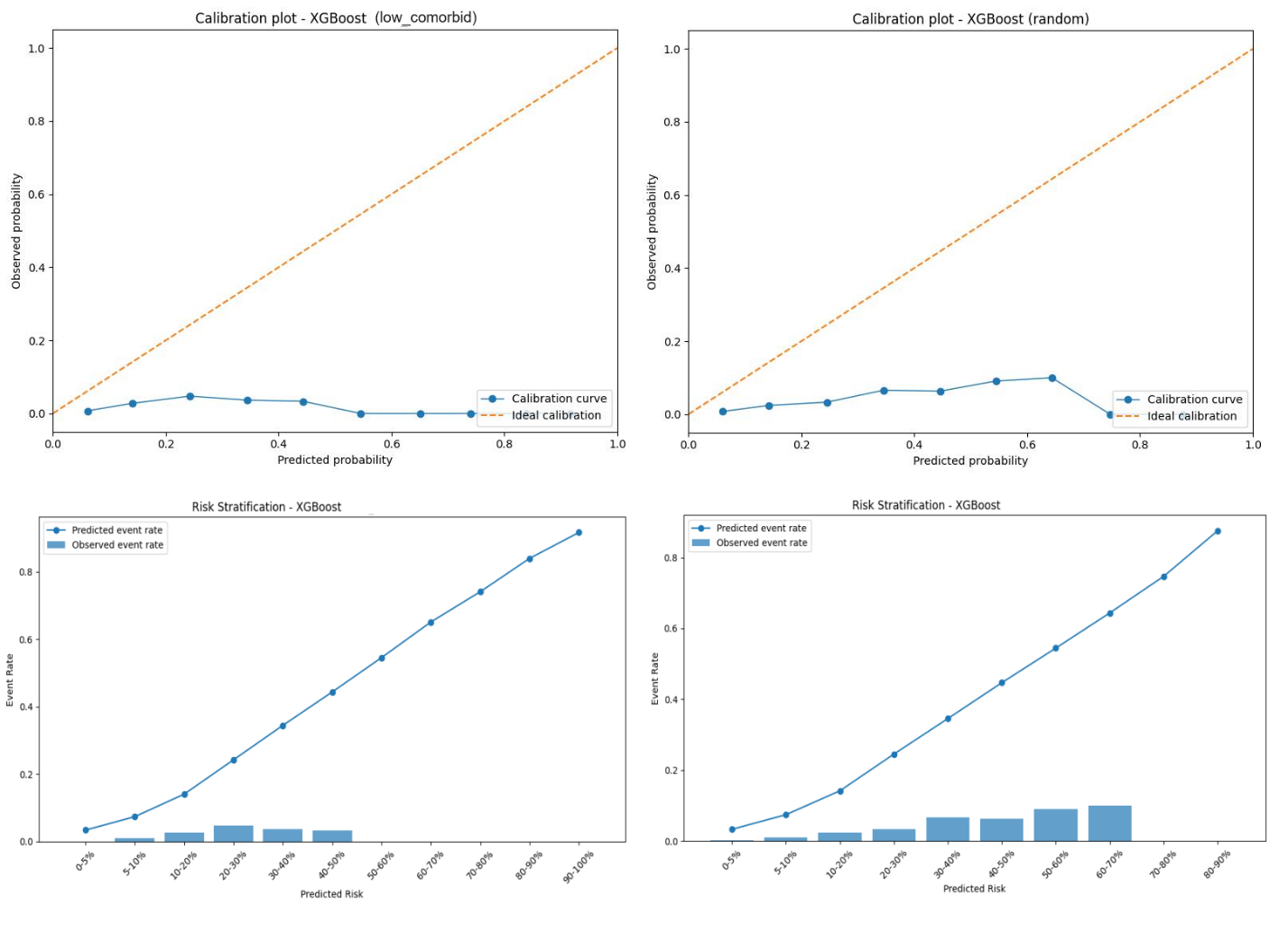

**Supplementary Figure 2.** Calibration plots and risk stratification plots for a) random forest model in the low comorbid and random test cohorts for prediction of 1-year bleeding risk
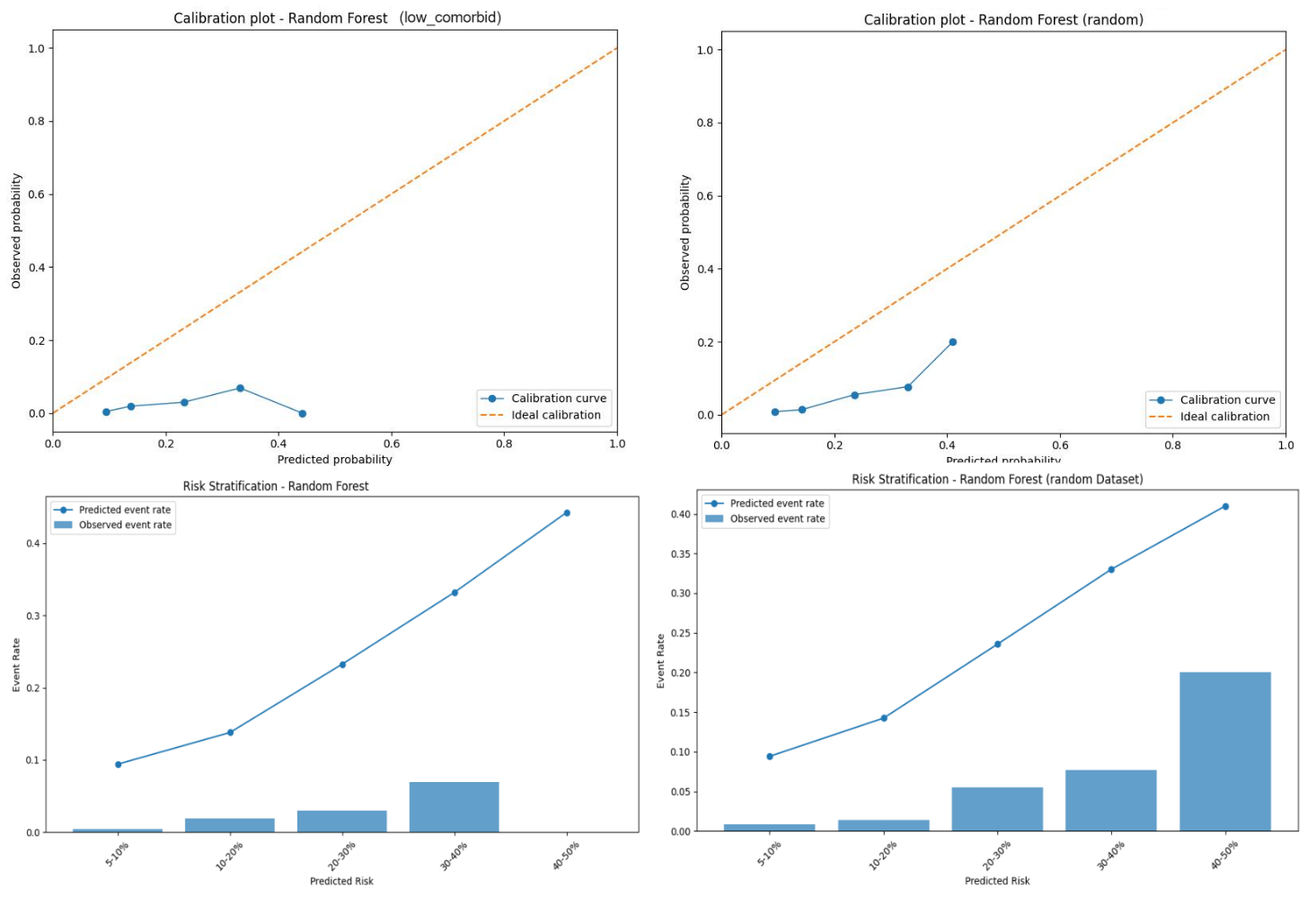

**Supplementary Figure 3.** Calibration plots and risk stratification plots for HASBLED risk score in the low comorbid and random test cohorts for prediction of 1-year bleeding risk.

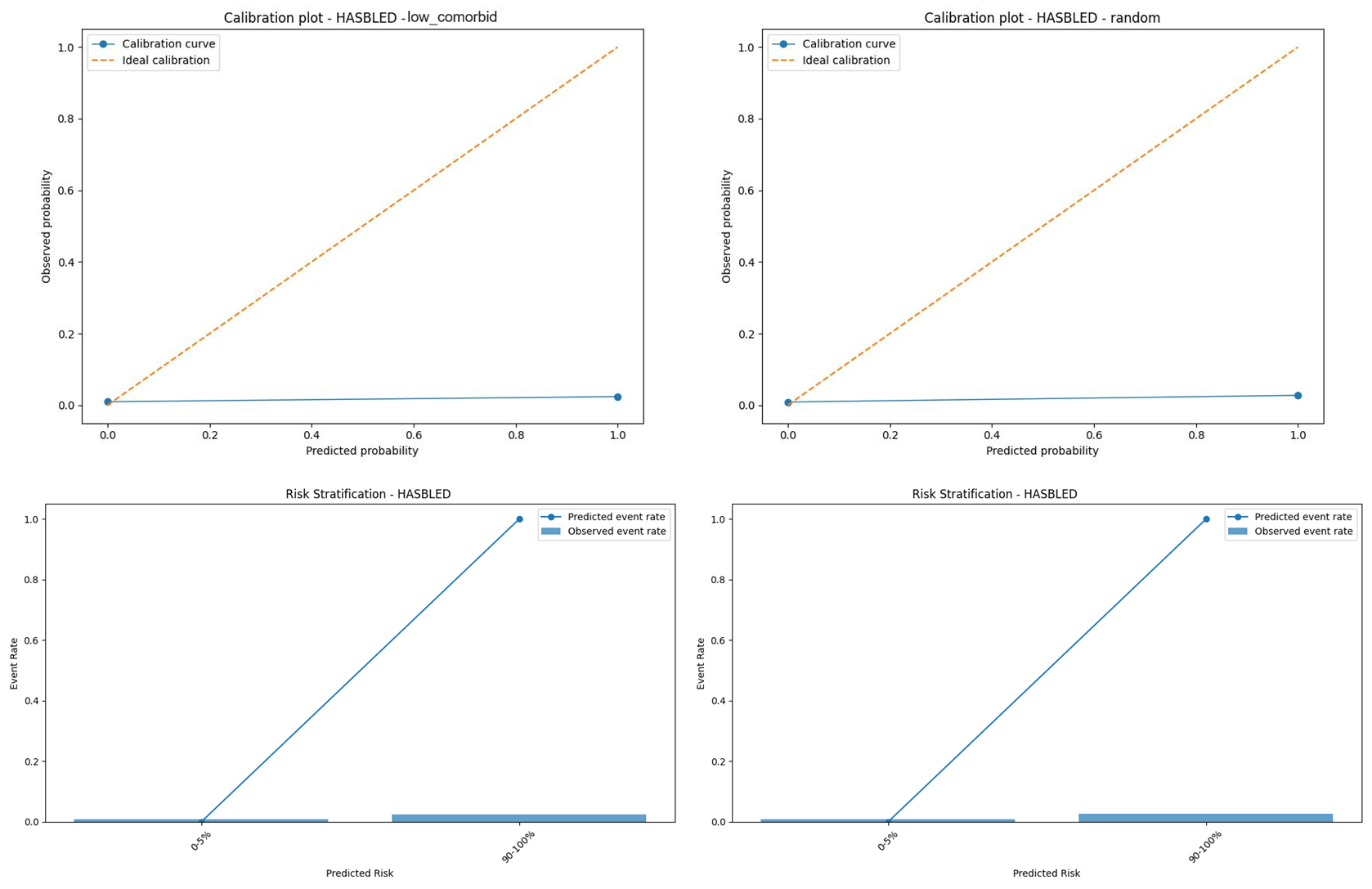

**Supplementary Figure 3.** Classification tree schema for prediction of major bleeding event at 1-year

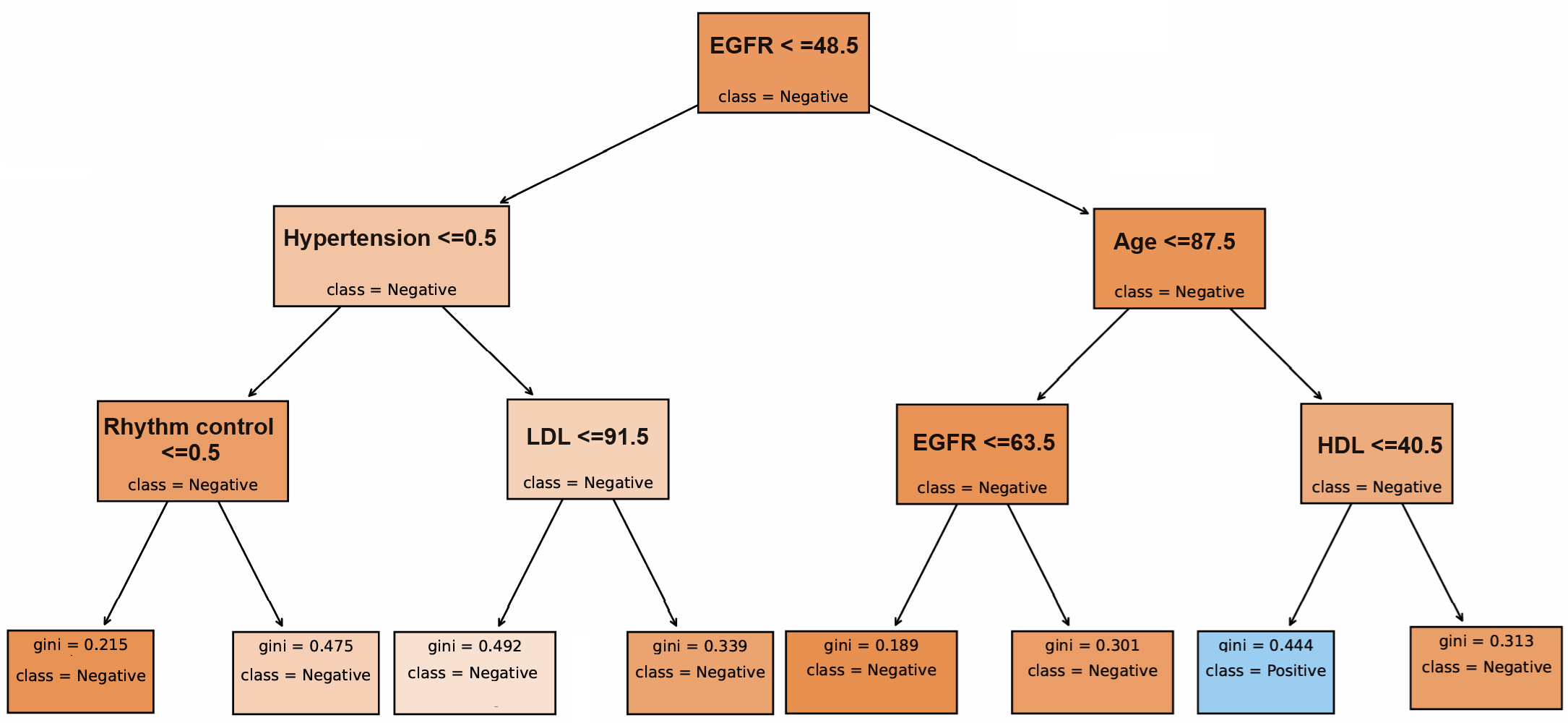
